# Microglial activation and tau burden predict cognitive decline in Alzheimer’s Disease

**DOI:** 10.1101/19011189

**Authors:** Maura Malpetti, Rogier A. Kievit, Luca Passamonti, P. Simon Jones, Kamen A. Tsvetanov, Timothy Rittman, Elijah Mak, Nicolas Nicastro, W. Richard Bevan-Jones, Li Su, Young T. Hong, Tim D. Fryer, Franklin I. Aigbirhio, John T. O’Brien, James B. Rowe

**Affiliations:** Department of Clinical Neurosciences, University of Cambridge, Cambridge, UK; MRC Cognition and Brain Sciences Unit, University of Cambridge, Cambridge, UK; Institute of Molecular Bioimaging and Physiology, National Research Council, Milano, Italy; Department of Psychiatry, University of Cambridge, Cambridge, UK; Department of Clinical Neurosciences, Geneva University Hospitals, Switzerland; Cambridge University Hospitals NHS Trust, Cambridge, UK

## Abstract

Tau pathology, neuroinflammation, and neurodegeneration are key aspects of Alzheimer’s disease. Understanding whether these features predict cognitive decline, alone or in combination, is crucial to develop new prognostic measures and enhanced stratification for clinical trials. Here, we studied how *baseline* assessments of *in vivo* tau pathology (measured by [^18^F]AV-1451 PET), neuroinflammation (indexed via [^11^C]PK11195 PET) and brain atrophy (derived from structural MRI) predicted *longitudinal* cognitive changes in patients with Alzheimer’s disease pathology. Twenty-six patients (n=12 with clinically probable Alzheimer’s dementia and n=14 with amyloid positive Mild Cognitive Impairment) and 29 healthy controls underwent baseline assessment with [^18^F]AV-1451 PET, [^11^C]PK11195 PET, and structural MRI. Cognition was examined annually over the subsequent 3 years using the revised Addenbrooke’s Cognitive Examination. Regional grey-matter volumes, [^18^F]AV-1451 and [^11^C]PK11195 binding were derived from fifteen temporo-parietal regions characteristically affected by Alzheimer’s disease pathology. A Principal Component Analysis (PCA) was used on each imaging modality separately, to identify the main spatial distributions of pathology. A Latent Growth Curve model was applied across the whole sample on longitudinal cognitive scores to estimate the rate of annual decline in each participant. We regressed the individuals’ estimated slope of cognitive decline on the neuroimaging components and examined univariable models with single-modality predictors, and a multi-modality model of prediction, to identify the independent and combined prognostic value of the different neuroimaging markers.

PCA identified a single component for the grey-matter atrophy, while two components were found for each PET ligand: one weighted to the anterior temporal lobe, and another weighted to posterior temporo-parietal regions. Across the whole-sample, the single-modality models indicated significant correlations between the slope of cognitive decline and the first component of each imaging modality. In patients, both stepwise backward elimination and Bayesian model selection revealed an optimal predictive model that included both components of [^18^F]AV-1451 and the first (i.e., anterior temporal) component for [^11^C]PK11195. However, the MRI-derived atrophy component and demographic variables were excluded from the optimal predictive model of cognitive decline. We conclude that temporo-parietal tau pathology and anterior temporal neuroinflammation predict cognitive decline in patients with symptomatic Alzheimer’s disease pathology. This indicates the added value of PET biomarkers in predicting cognitive decline in Alzheimer’s disease, over and above MRI measures of brain atrophy and demographic data. Our findings also support the strategy for targeting tau and neuroinflammation in disease-modifying therapy against Alzheimer’s Disease.

## Introduction

The pathological hallmarks of Alzheimer’s disease are tau neurofibrillary tangles (NFT) and amyloid-β plaques, but neuroinflammation has also emerged as a key process in Alzheimer’s disease and other neurodegenerative disorders (Pasqualetti *et al*., 2015; Ransohoff, 2016; Schain and Kreisl, 2017). However, the differential role of these pathologies in predicting clinical progression of Alzheimer’s disease remains to be ascertained. This represents a critical step to develop new prognostic markers and test the effect of novel disease modifying therapies that target different pathologies in Alzheimer’s disease.

The aggregation of misfolded tau protein plays a fundamental role in synaptic dysfunction and neuronal loss, and correlates with clinical severity in the Alzheimer’s disease clinical spectrum (Nelson *et al*., 2012; Spires-Jones and Hyman, 2014). A significant presence of amyloid-β plaques is also indicative of likely cognitive decline in mid- and later-life, although the association of both neurodegeneration and cognitive impairment has been found stronger with the distribution and burden of NFTs than it is for neuritic plaques (Nelson *et al*., 2012; Spires-Jones and Hyman, 2014). Microglia activation and neuroinflammation represent a third key determinant in the etio-pathogenesis of Alzheimer’s disease and in its progression (Heneka *et al*., 2015; Mhatre *et al*., 2015; Calsolaro and Edison, 2016), independently or synergistically with tau and amyloid pathology.

Each of these processes can now be quantified and localised indirectly *in vivo* using brain imaging, such as PET imaging with radioligands targeting tau pathology, amyloid burden, and microglial activation (see Chandra *et al*., 2019 for a review). The PET ligand [^18^F]AV-1451 is sensitive to cortical tau accumulation in Alzheimer’s disease, and has high affinity for the characteristic paired helical tau filaments (Xia *et al*., 2013; Marquié *et al*., 2015; Lowe *et al*., 2016). [^18^F]AV-1451 PET studies have shown marked tau accumulation in the entorhinal cortex in patients with mild cognitive impairment (MCI) which extends to temporo-parietal regions in Alzheimer’s disease (Hall *et al*., 2017). [^18^F]AV-1451 bindings also correlates with Braak staging of neurofibrillary tau (Schwarz *et al*., 2016; Schöll *et al*., 2018), and post-mortem patterns of Alzheimer’s disease pathology (Sander *et al*., 2016). This is also in keeping with evidence that tau deposition is evident as a continuum from normal aging through MCI to Alzheimer’s dementia (Schöll *et al*., 2018), and correlates with cognitive impairment (Brier *et al*., 2016, Cho *et al*., 2016*b*; Johnson *et al*., 2016; Ossenkoppele *et al*., 2016; Pontecorvo *et al*., 2017). In addition, PET imaging supported the previous evidence on a stronger association of cognitive deficits with tau burden than with amyloid-β (Brier *et al*., 2016; Johnson *et al*., 2016).

The PET ligand [^11^C]PK11195 is a well-established marker for microglial activation in Alzheimer’s disease via its binding to the 18-kDa translocator protein (TSPO), a mitochondrial membrane protein which is overexpressed in activated microglia (Scarf and Kassiou, 2011). In Alzheimer’s disease, [^11^C]PK11195 shows high binding in temporo-parietal regions and cingulate cortex (Stefaniak and O’Brien, 2015). Higher levels of neuroinflammation in these regions is also negatively associated with cognitive performance in MCI and Alzheimer’s dementia (Edison *et al*., 2008; Okello *et al*., 2009, Fan *et al*., 2015*a*; Passamonti *et al*., 2018, 2019) although they do not correlate well with amyloid burden (Yokokura *et al*., 2011), suggesting an independent role of microglia activation in mediating cognitive deficit.

There are extensive data on atrophy in Alzheimer’s Disease, measured in terms of volume loss *in vivo* by MRI, at MCI and dementia stages of progressive Alzheimer’s Disease pathology. MRI measures, for example of medial temporal lobe volumes, correlates with disease severity, and are predictive of future conversion from MCI to Alzheimer’s Disease (Frisoni *et al*., 2010; Leung *et al*., 2013; Jack *et al*., 2015). However, cell loss and atrophy are a relatively late feature in a cascade of pathology, and it is not clear how MRI compares with measures of molecular pathology as a prognostic marker, especially in view of marked age-related structural changes (Raz *et al*., 2005; Walhovd *et al*., 2011).

In this study, we use baseline *in vivo* measures of tau pathology, microglia activation, and brain atrophy to predict the rate of cognitive decline in patients with Alzheimer’s disease pathology, ranging from MCI with biomarker evidence of amyloid pathology to clinically probable Alzheimer’s disease. Our hypothesis was that the PET biomarkers of tau pathology and neuroinflammation are strong predictors of cognitive impairment and decline, and that whereas MRI may be predictive in isolation, the prognostic information of MRI is better captured by direct measures of molecular pathology using PET (Bejanin *et al*., 2017; Mattsson *et al*., 2019). This hypothesis builds on the evidence that tau burden relates to age-related cognitive decline (Schöll *et al*., 2016; Aschenbrenner *et al*., 2018; Maass *et al*., 2018), and progression of dementia over 6 to 18 months in patients with Alzheimer’s disease (Koychev *et al*., 2017; Pontecorvo *et al*., 2019). In contrast to past studies that have assessed the relationship between longitudinal PET markers and clinical changes in Alzheimer’s Disease (Fan *et al*., 2015*b*, 2017; Chiotis *et al*., 2018; Jack *et al*., 2018; Southekal *et al*., 2018; Cho *et al*., 2019), we study how a multi-modal and cross-sectional assessment of distinct pathologies is able to predict longitudinal decline in Alzheimer’s Disease, examining the individual or combined prognostic contribution of tau pathology, neuroinflammation, and brain atrophy in predicting cognitive decline.

A better characterisation of the factors predicting decline in Alzheimer’s disease will help to develop enhanced prognostic and outcome measures for clinical trials targeting more than one pathology. Although previous findings support the use of MRI and PET imaging in the diagnosis and monitoring of disease progression, the prognostic value of these *in vivo* measures and their combined effect in predicting clinical decline in Alzheimer’s disease remains undetermined. Previous studies which have evaluated the predictive values of neuroimaging markers in Alzheimer’s disease have typically assessed different neuroimaging modalities in isolation rather than exploiting the mechanistic and prognostic values that is offered by multi-modal neuroimaging. We therefore assessed the independent and combined predictive effects of baseline neuroimaging biomarkers for tau pathology ([^18^F]AV-1451 PET), neuroinflammation ([^11^C]PK11195 PET) and brain atrophy (structural MRI) on longitudinal cognitive changes over a period of three years in the clinical spectrum of Alzheimer’s disease.

Focussing therefore on temporo-parietal regions, we predicted: 1) a significant association between baseline measures of each neuroimaging technique and longitudinal decline in cognition, assessed separately for each modality; 2) partially independent and additive effects of MRI and PET measures on cognitive decline, assessed with all modalities in a single multivariate model. Moreover, we predicted that the molecular markers of baseline tau and neuroinflammation PET could be more informative than structural MRI on longitudinal cognitive deterioration in Alzheimer’s disease.

## Material and methods

### Participants

We recruited 26 patients: twelve with a clinical diagnosis of probable Alzheimer’s dementia and 14 with amnestic MCI and a positive amyloid PET scan as biomarker of Alzheimer’s Disease (Klunk *et al*., 2004). Probable Alzheimer’s dementia was diagnosed according to the National Institute on Aging-Alzheimer’s Association guidelines (McKhann *et al*., 2011) and confirmed in all patients during follow-up. Given the long-term and intensive nature of the longitudinal project, all patients at baseline had >12/30 on the Mini-Mental State Examination (MMSE) to be eligible to the study. MCI patients had MMSE score > 24/30, and memory impairment not ascribable another diagnosis (Albert *et al*., 2011). We also included 29 healthy controls with MMSE >26/30, absence of memory symptoms, no signs of dementia, or any other significant medical illnesses.

All gave informed consent according to the Declaration of Helsinki. The NIMROD protocol (Neuroimaging of Inflammation in Memory and Related Other Disorders, (Bevan-Jones *et al*., 2017)) was approved by the NIHR National Research Ethic Service Committee and East of England (Cambridge Central).

During the first visit, demographic information and medical history were collected. All participants underwent baseline neuropsychological assessment, MRI scan and one, two or three PET scans depending on the group. The clinical examination and neuropsychological battery were repeated annually for three follow-up visits (see Bevan-Jones et al., 2017 for more details). The revised Addenbrooke’s Cognitive Examination (ACE-R) (Mioshi *et al*., 2006) was used to assess the cognitive performance at each visit.

### Imaging data acquisition and pre-processing

At the baseline, all subjects underwent 3T MRI performed on a Siemens Magnetom Tim Trio or Verio scanner (Siemens Healthineers, Erlangen, Germany). MCI and AD subjects had both [^18^F]AV-1451 PET and [^11^C]PK11195 PET, while, to minimise radiation exposure to healthy people, control subjects were divided into two groups: 14 underwent [^18^F]AV-1451 PET, while another 15 underwent [^11^C]PK11195 PET. PET scanning was undertaken on a GE Advance PET scanner (GE Healthcare, Waukesha, USA) and a GE Discovery 690 PET/CT (see (Passamonti *et al*., 2017, 2018) for more details). Patients with MCI also underwent 40-70 minutes post-injection [^11^C]PiB PET to quantify the density of fibrillar Aβ deposits for classification of Aβ status. [^11^C]PiB scans were classified as positive if the average standardized uptake value ratio (SUVR) across the cortex using a cerebellar grey matter reference region was > 1.5. MCI subjects with positive Aβ status were included in this study.

T1-weighted MRI was used for tissue class segmentation into grey matter, white matter and cerebrospinal fluid (CSF) using SPM12. Segmented images were used to determine regional grey matter, white matter and CSF volumes, and to calculate brain volume (grey + white matter) and total intracranial volume (TIV = grey matter + white matter + CSF) in each participant. Grey and white matter segments from 33 representative subjects were imported to create an unbiased template (11 controls, 11 patients with mild cognitive impairment and 11 patients with Alzheimer’s Dementia, matched for age and sex across the groups) using the DARTEL pipeline in SPM12. All other images were subsequently warped and aligned to the template. The group template was warped to the MNI152 template using *‘Population to ICBM’* function. The deformations from this transformation were combined with the flow fields of each individual and inverted. Eighty-three cortical and subcortical regions of interest (ROIs) were defined using a version of the Hammers atlas (Gousias *et al*., 2008) modified to include brainstem parcellation and the cerebellar dentate nucleus. All segmented images were modulated and warped to the modified Hammers atlas in MNI space. Individual regional grey matter volumes were then extracted from the spatially normalised and modulated grey matter segments, using the *‘spm summarise atlas’* function.

For each subject, the aligned PET image series for each scan was rigidly co-registered to the T1-weighted MRI image. Prior to kinetic modeling regional PET data were corrected for CSF contamination by dividing by the mean region grey plus white matter fraction determined from SPM tissue probability maps smoothed to PET spatial resolution. For [^11^C]PK11195, supervised cluster analysis was used to determine the reference tissue time-activity curve and non-displaceable binding potential (BP_ND_) was calculated in each ROI using a simplified reference tissue model that includes correction for vascular binding (Yaqub *et al*., 2012). For [^18^F]AV-1451, BP_ND_ was assessed in each ROI with the simplified reference tissue model (Gunn *et al*., 1997) using superior cerebellar cortex grey matter as the reference region. For more details about data acquisition and pre-processing steps see Passamonti et al. (Passamonti *et al*., 2017, 2018).

### Statistical analyses

#### Descriptive statistics

Continuous variables (age, education, ACE-R) were compared between groups with an independent-samples t-test, and categorical variables (sex) with the Chi-square test. The effect size of each t-test comparison was computed to quantify differences between the two groups (Cohen’s d > 0.8, valuable difference). ACE-R scores at follow-up were annualised to the nearest whole year.

#### Principal components analysis

The number of ROIs was reduced from 83 to 15, by a) combining left and right regional values, as in our previous studies on Alzheimer’s disease (Passamonti *et al*., 2017, 2018), and b) focussing on *a priori* regions related to Alzheimer’s disease pathology (see Supplementary Table 1). Considering the literature showing early neurodegeneration, tau pathology, and neuroinflammation in temporal and parietal brain regions in Alzheimer’s disease (i.e., temporo-parietal regions) we focused our analyses on those areas. Regional grey matter volume used the sum of the grey matter volumes in the corresponding left and right hemisphere ROIs, corrected for TIV. For both [^11^C]PK11195 and [^18^F]AV-1451, a volume-weighted mean of left and right regional BP_ND_ values was calculated. Values determined for the 15 bilateral ROIs from each imaging dataset were included in three principal component analyses (PCAs), run separately for grey matter volumes, [^11^C]PK11195 and [^18^F]AV-1451 BP_ND_ values. This reduces dimensionality and the problem of multiple comparisons, identifying a limited number of components that best explain the data variance. We applied an orthogonal Varimax rotation to maximize interpretability and specificity of the resulting components. We retained components with eigenvalues >1. To test whether correction for CSF affected the PCA results, we applied the same analyses on [^18^F]AV-1451 PET and [^11^C]PK11195 regional data not corrected for CSF partial volume.

**Table 1.**
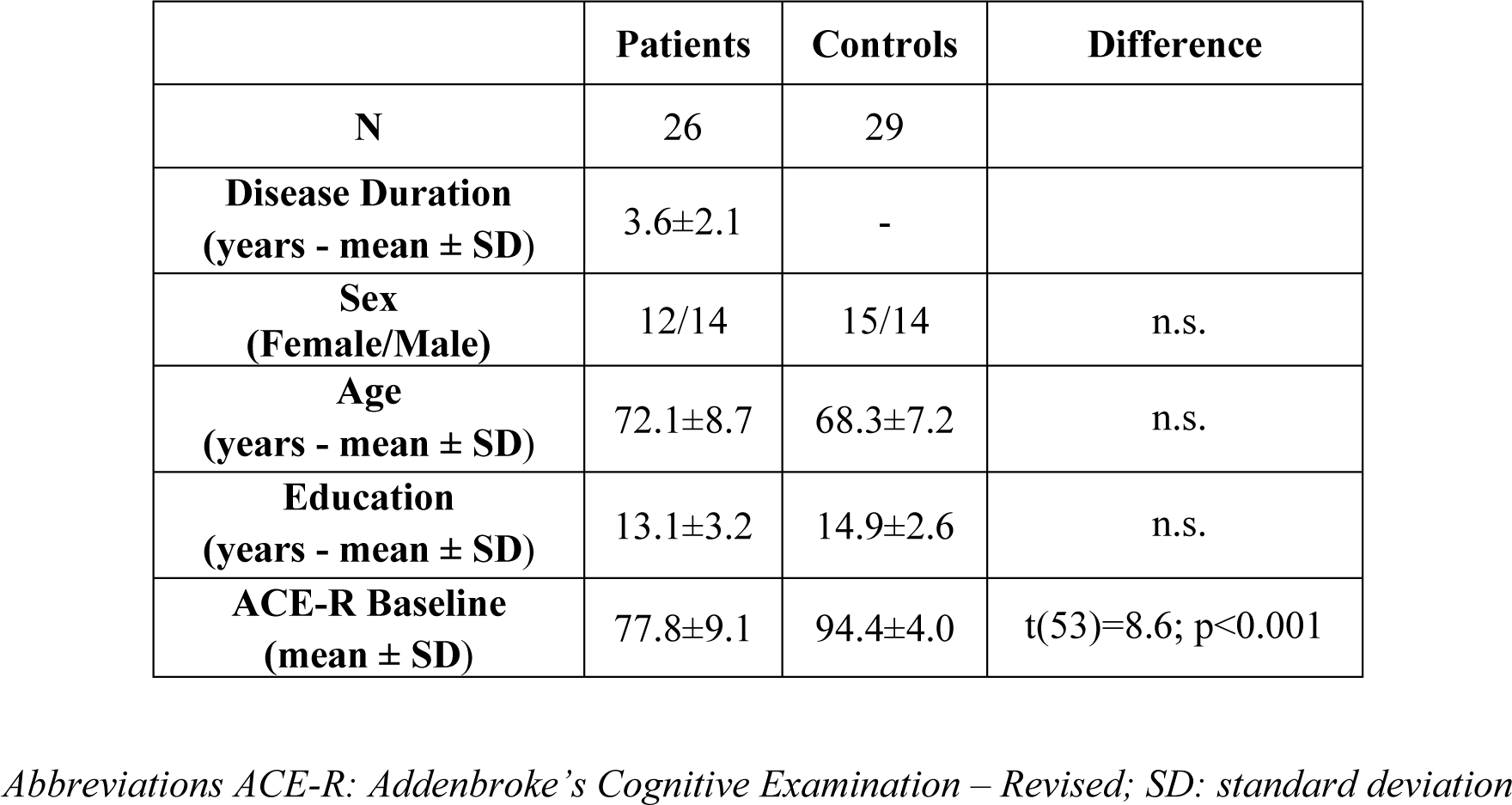
Demographic and clinical characteristics for the patient and control groups. (n.s.= non-significant difference between patients and controls, defined as p-value > 0.05 or Choen’s d < 0.8).

|  | **Patients** | **Controls** | **Difference** |
| --- | --- | --- | --- |
| **N** | 26 | 29 |  |
| **Disease Duration**<br>**(years - mean ± SD)** | 3.6±2.1 | - | |
| **Sex**<br>**(Female/Male)** | 12/14 | 15/14 | n.s. |
| **Age**<br>**(years - mean ± SD)** | 72.1±8.7 | 68.3±7.2 | n.s. |
| **Education**<br>**(years - mean ± SD)** | 13.1±3.2 | 14.9±2.6 | n.s. |
| **ACE-R Baseline**<br>**(mean ± SD)** | 77.8±9.1 | 94.4±4.0 | t(53)=8.6; p<0.001 |

*Abbreviations ACE-R: Addenbroke's Cognitive Examination – Revised; SD: standard deviation*

The individual component scores were corrected for the time interval in months between the scan and the baseline cognitive assessment. Mean and standard deviation of the time interval between the imaging scans and the baseline cognitive assessment were: 1.75 ± 2.50 months for MRI, 7.18 ± 5.68 months for [^18^F]AV-1451 PET and 6.12 ± 9.07 months for [^11^C]PK11195 PET. The residuals extracted for each component were included in a multiple regression on cognitive decline as independent variables.

#### Latent Growth Curve Model for cognitive data

An univariable LGCM was fitted on longitudinal ACE-R scores across all subjects (n=55), to obtain the (i) intercept; (ii) slope, quantifying the rate of change and its form (i.e. linear or nonlinear); (iii) the relation between intercept and slope. The estimated parameters are based on the individuals’ trajectory, indicating average change and individual difference. Covariates can be added to the model to assess their associations with both intercept and slope. Three time points and 5-10 cases per parameter are required for a standard LGCM (Bentler and Chou, 1987; Newsom, 2015). LGCM was implemented in Lavaan software (Rosseel, 2012) using full information maximum likelihood estimation with robust standard errors for missingness and non-normality. We considered four indices of good model fit (Schermelleh-Engel *et al*., 2003): 1) the chi-square test with the p-value (good fit: > 0.05), 2) the root-mean-square error of approximation (RMSEA, acceptable fit: < 0.08, good fit: < 0.05), 3) the comparative fit index (CFI, acceptable fit: 0.95–0.97, good fit: > 0.97), and 4) the standardized root mean-square residual (SRMR, acceptable fit: 0.05–0.10, good fit: < 0.05). From the model fitting, variables “intercept” and “slope” were created extracting the individual estimated values for each subject in the model. T-tests and analysis of variance tested for group differences in initial cognitive performance and annual change.

#### One-step prediction procedure: Latent Growth Curve Model with predictors

First, across all subjects, we tested the predictive value of each imaging method on cognitive decline, applying five LGCMs with each scan-specific component’s values (corrected for months from the baseline) as predictor of cognitive intercept and slope. The models were tested separately for MRI (n=55), [^11^C]PK11195 PET (n=41) and [^18^F]AV-1451 PET (n=40) components. Second, in patients only (n=26), the individual scores of all imaging components were included as predictors in the LGCM on longitudinal ACE-R scores, to test the combined predictive effect of all imaging modalities.

The one step procedure would be the ideal approach to answer our research questions, however, this leads to model estimation challenges in a cohort with a modest sample size. For this reason, we next applied a two-step prediction procedure: 1) extracting individual slope values from the initial LGCM for cognitive data across all the population, and 2) including these values as dependent variable in linear regression models with brain imaging components as predictors. We present both frequentist and Bayesian analyses to ensure inferential robustness and allowing us to quantify evidence in favour of the null hypothesis.

#### Two-step frequentist prediction procedure: linear regression models on LGCM estimated parameters

First, across all subjects, the residual values of each scan-specific PCA component (corrected for months from the baseline) were included as single predictors in separated univariable linear regression models with the individual slope values extracted from the initial LGCM as dependent variable. The significance level was set at p<0.01 corrected for multiple comparisons with the Bonferroni correction (p=0.05/5). Next, the individual scores of the imaging methods’ components were included as independent variables in a multivariable regression analysis on patients alone (N=26), who underwent all three imaging scans. This model was fit to examine the individual, as well as combined, ability to explain variance in cognitive decline using brain marker components as well s age, education, and sex as independent variables. The model used stepwise backward selection (entry criterion p=0.05 and elimination criterion p=0.1). In a supplementary analysis, we also applied a “reduced” multivariable linear regression analysis with slope as dependent variable and only the first component of each imaging method as predictors to test that the different number of components between MRI and PET did not affect the estimation. Given the challenges of stepwise model selection, and the limitations of sample size to utilize more optimal methods (e.g. regularized model fitting), we ran the analysis using Bayesian methodology, to ensure inferential robustness of our findings and confidence in null results.

#### Two-step Bayesian prediction procedure: linear regression models on LGCM estimated parameters

Finally, we applied a Bayesian multiple regression analysis with brain components and demographic variables as predictors, and the estimated slope values as dependent variable. This approach was used to test whether there was evidence for the absence of independent variables’ effect for those components excluded from the final models (as opposed to frequentist type II error). In the model comparisons, we considered as final model the one with the highest Bayes Factor compared to the null model (BF_10_). Then, we used a reduced Bayesian linear regression, mirroring the reduced model applied with the frequentist approach, which included only the first component of each imaging method as predictors of slope.

See Supplementary Fig. 1 for a schematic representation of statistical analyses with one-step and two-step prediction procedures.

**Figure 1.**
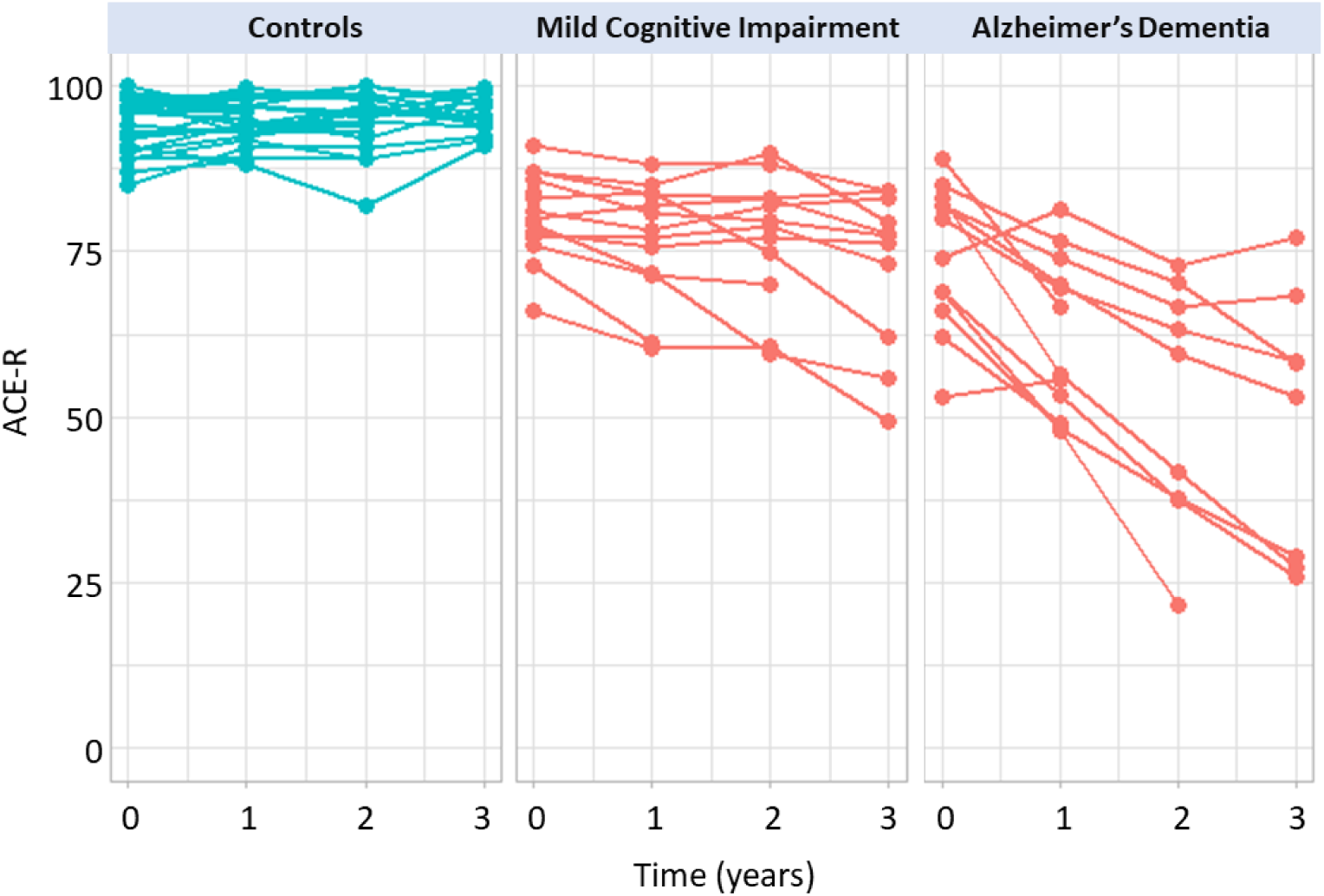
Longitudinal cognitive changes in patients and controls, as measured by revised Addenbrooke’s Cognitive Examination (ACE-R). Points represent annualised ACE-R scores at baseline, 1-year, 2-years and 3-years follow-up for each subject in control (blue) and patient (red) groups.

### Data availability

Anonymized data may be shared by request to the senior author from a qualified investigator for non-commercial use (data sharing is subject to participants’ prior consent to data sharing).

## Results

### Descriptive statistics

Significant differences between patient and control groups were found for education (t(53)=2.4, p=0.02, Cohen’s d=0.64) and ACE-R scores (t(53)=8.6, p<0.001, Cohen’s d=2.37), however, the effect size of the education difference was negligible (d < 0.08). There were not significant group differences in age (t(53)=-1.7, p=0.09, Cohen’s d=-0.47) and sex (χ2(1)=0.17, p=0.68) (Table 1). Individual ACE-R scores at baseline and at each follow-up are shown in Fig. 1.

### Principal component analysis of Grey Matter Volumes, [^18^F]AV-1451 BP_ND_ and [^11^C]PK11195 BP_ND_

For grey-matter volumes, the PCA on the pre-selected 15 AD-specific cortical regions identified only one component that encompassed all the temporo-parietal regions and explained 74% of the variance (Fig. 2, left panel). Two principal components were detected for [^18^F]AV-1451 BP_ND_ data, explaining 91% of the total variance (83% first component; 8% second component). The first component was loaded onto the posterior temporal and parietal regions, while the second component was weighted to the anterior temporal lobe, amygdala, insula and hippocampus (Fig. 2, middle panel). For [^11^C]PK11195 BP_ND_ data, two principal components were identified, and these explained together the 76% of data variance (56% for the first component; 20% for the second component). The first component involved anterior and medial temporal lobe, while the second component was mainly loaded onto the posterior temporo-parietal regions and insula (Fig. 2, right panel). Using PET data without CSF correction yielded qualitatively similar results.

**Figure 2.**
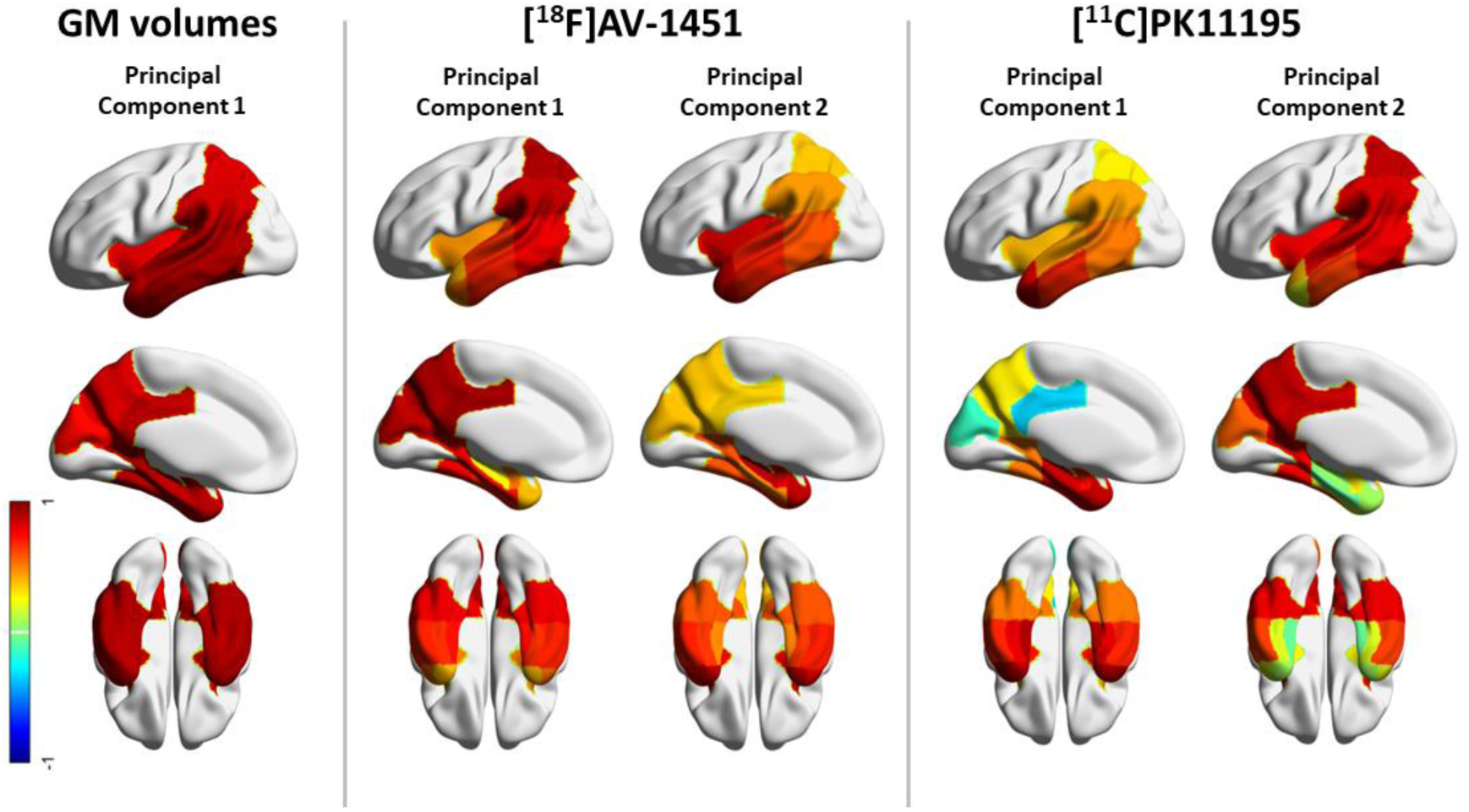
Regional weights of the structural MRI component (left), and rotated regional weights of [^18^F]AV-1451 components (middle) and the [^11^C]PK11195 components (right). Components were identified applying three independent principal component analyses on 15 temporo-parietal regions. For structural MRI, regional grey matter (GM) volumes were included in the analysis, while for each PET tracer, the binding potential values in those regions were considered, separately for each modality.

### Subject-specific rate of cognitive decline

The LGCM of longitudinal ACE-R scores fitted the data adequately (χ2(8)=10.93, p=0.206; RMSEA=0.09 [0.00 – 0.21], CFI=0.99, SRMR=0.04). The mean of the intercept was 86.40 (Standard Error (SE)=1.44, z-value=60.02, fully standardized estimate (Std Est) =8.28, p<0.001) and average cognition declined over time (slope, estimate (est)=-3.01, SE=0.80, z-value=-3.75, Std Est=-0.54, p<0.001). The intercept significantly co-varied with the slope (est=38.51, SE=9.24, z-value=4.17, Std Est (correlation)=0.67, p<0.001, such that individuals with higher (better) baseline performance showed less steep decline. As expected, patients significantly differed from controls in their intercept (t(53)=9.39, p<0.001) and slope (t(53)=6.42, p<0.001) indicating a faster and more severe cognitive decline (Fig. 3). Across three groups, ANOVA confirmed group differences in the intercept (F(2)=63.44, p<0.001; mean (SD) for: HC=94.18 (3.27); MCI+=81.25 (6.17); AD=73.60 (8.96)) and slope (F(2)=53.74, p<0.001; mean (SD) for: HC=0.40 (0.82); MCI+=-3.56 (3.08); AD=-10.62 (5.71)), with post-hoc confirmation of differences between each pair of groups (all p<0.005).

**Figure 3.**
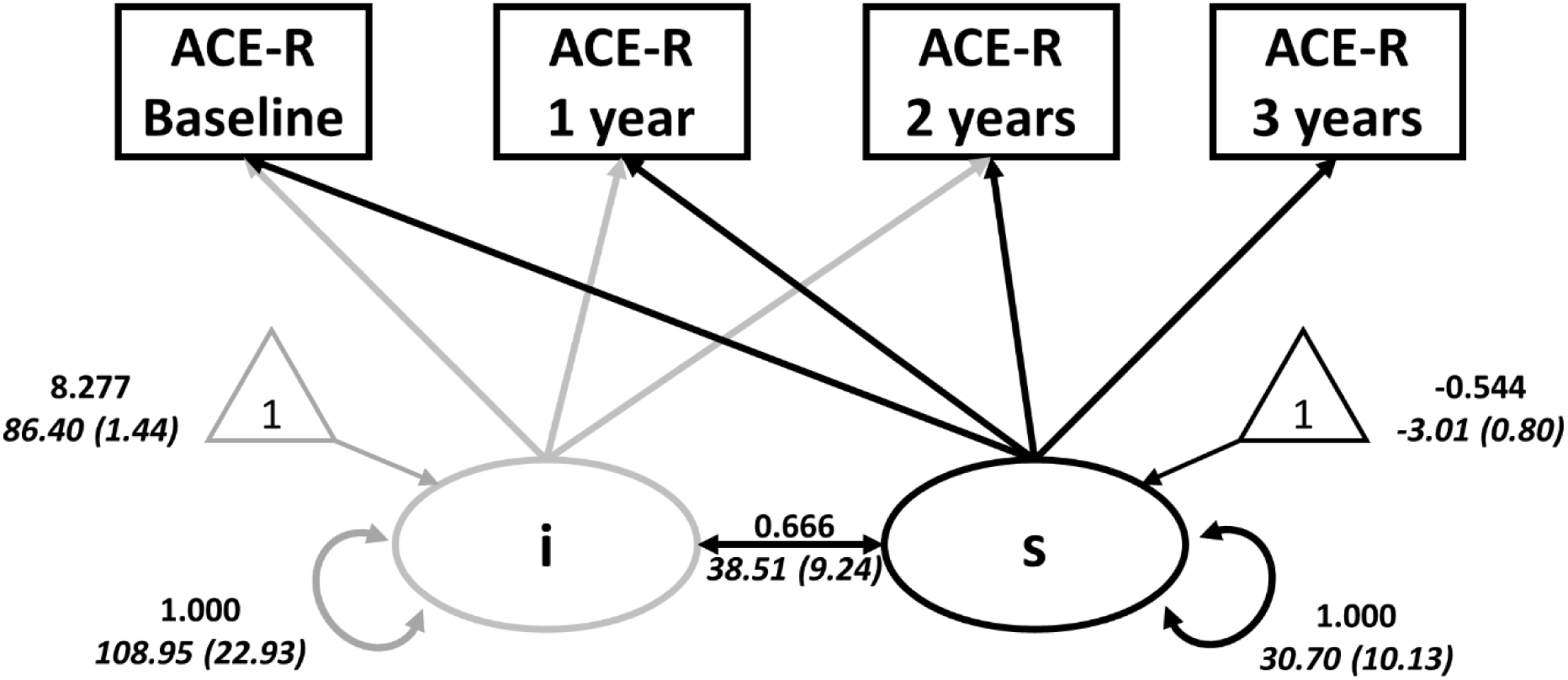
Latent growth curve model to test the initial values (intercept – “i”) and longitudinal changes (slope – “s”) in scores of revised Addenbrooke’s Cognitive Examination (ACE-R) across all sample. Circles indicate latent variables, rectangles indicate observed variables, and triangles denote intercepts (means). Thick single-headed arrows indicate regressions while thick double-headed arrows indicate variance and covariance (grey for intercept and black for slope). Values in Roman are standardized parameter estimates, and values in italics are unstandardized parameter estimates (with standard errors in parentheses). The annual rate of change was positively associated with performance at baseline (lower initial cognitive scores were associated with a higher annual rate of cognitive changes).

### One-step prediction Latent Growth Curve Model with predictors

The LGCM including MRI fitted the data marginally well (χ2(10)=18.33, p-value=0.05, RMSEA=0.13 [0.01-0.23], CFI=0.98, SRMR=0.03). Individual differences in the summary brain measure were strongly and positively associated with both slope (path Std Est=0.58, p<0.001) and intercept (path Std Est=0.67, p<0.001). This suggested that individuals with greater grey matter volumes showed better baseline performance, and slower longitudinal decline, than those with smaller volumes.

The LGCM of the posterior [^18^F]AV-1451 component fitted the data adequately (χ2(10)=16.30, p-value=0.09, RMSEA=0.12 [0.00-0.22], CFI=0.98, SRMR=0.03). Here too, both the slope (path Std Est=-0.62, p=0.001) and intercept (path Std Est=-0.53, p<0.001) were strongly governed by individual differences in the first component. In contrast, in the model with only the anterior [^18^F]AV-1451 component (χ2(10)=21.75, p-value=0.01, RMSEA=0.17 [0.07-0.27], CFI=0.96, SRMR=0.05), there was no association between the scores on the neural component and either the intercept (path Std Est=-0.12, p=0.431) or the slope (path Std Est=-0.39, p=0.057).

Finally, the LGCM with the anterior [^11^C]PK11195 component fitted the data adequately (χ2(10)=16.32, p-value=0.09, RMSEA=0.13 [0.00-0.23], CFI=0.97, SRMR=0.02). Individual differences in the [^11^C]PK11195 component governed both slope (Std Est=-0.51, p=0.002) and intercept (Std Est=-0.43, p<0.001) correlated with the component #1. In the model with the posterior [^11^C]PK11195 component as regressor (χ2(10)=9.33, p-value=0.50, RMSEA=0.00 [0.00-0.17], CFI=1.00, SRMR=0.03), the slope resulted significantly correlated with the component (Std Est=-0.45, p=0.009), but not the intercept (Std Est=-0.010, p=0.951).

In patients, an LGCM including the components of all three imaging methods did not fit well (χ2(18)=34.76, p-value=0.01, RMSEA=0.17 [0.08 – 0.26], CFI=0.92, SRMR=0.04). With this caveat, cognitive decline (slope) was predicted by baseline posterior [^18^F]AV-1451 (path Std Est=-0.49, p=0.025) and anterior [^11^C]PK11195 (path Std Est=-0.40, p=0.017) components’ scores, but not the posterior [^11^C]PK11195, the MRI (path Std Est=0.10, p=0.52) or the anterior [^18^F]AV-1451 (Std Est=-0.22, p=0.23) components.

### Two-step prediction: linear regression

Across all subjects, the rate of cognitive decline (slope from LGCM) was significantly associated with: 1) the MRI weighting (Std Beta=0.61, p<0.001); 2) the posterior [^18^F]AV-1451 (Std Beta=-0.60, p<0.001); 3) and anterior [^11^C]PK11195 (R=-0.47, p=0.002). All these results survived Bonferroni’s correction. Correlations of slope with the anterior [^18^F]AV-1451 (Std Beta=-0.36, p=0.022), and the posterior [^11^C]PK11195 (Std Beta=-0.39, p=0.012) did not survive correction for multiple comparisons (p<0.01). See Fig. 4 for a graphical representation of the significant associations between individual scores (x axis) of imaging-specific principal components and slope in ACE-R scores (y axis) extracted by LGCM. Model summary and coefficients for all univariable models with slope as dependent variable are reported in Table 2.

**Table 2.** Results for the univariable regression models on slope across all population (p=uncorrected p-values, *= Bonferroni corrected, significance threshold p<0.01).

| Model |  | Estimate | Std Error | Std Beta | t value | p | Adj R<sup>2</sup><br>(std err) | F | p |
| --- | --- | --- | --- | --- | --- | --- | --- | --- | --- |
| **MRI**<br>(N=55) | **(Intercept)** | -3.01 | 0.58 |  | -5.23 | 0.000 | 0.358<br>(4.27) | 31.18 | <0.001* |
|  | **MRI component** | 3.48 | 0.63 | 0.61 | 5.58 | 0.000 |  |  |  |
| **AV 1**<br>(N=40) | **(Intercept)** | -4.31 | 0.73 |  | -5.88 | 0.000 | 0.341<br>(4.64) | 21.22 | <0.001* |
|  | **AV component 1** | -3.43 | 0.74 | -0.60 | -4.61 | 0.000 |  |  |  |
| **AV 2**<br>(N=40) | **(Intercept)** | -4.31 | 0.85 |  | -5.05 | 0.000 | 0.108<br>(5.40) | 5.72 | 0.022 |
|  | **AV component 2** | -2.08 | 0.87 | -0.36 | -2.39 | 0.022 |  |  |  |
| **PK 1**<br>(N=41) | **(Intercept)** | -4.15 | 0.80 |  | -5.19 | 0.000 | 0.204<br>(5.13) | 11.26 | 0.002* |
|  | **PK component 1** | -2.72 | 0.81 | -0.47 | -3.36 | 0.002 |  |  |  |
| **PK 2**<br>(N=41) | **(Intercept)** | -4.15 | 0.84 |  | -4.96 | 0.000 | 0.128<br>(5.36) | 6.87 | 0.012 |
|  | **PK component 2** | -2.36 | 0.90 | -0.39 | -2.62 | 0.012 |  |  |  |

Abbreviations AV: [<sup>18</sup>F]AV-1451; PK: [<sup>11</sup>C]PK11195; Std: standard; Adj: adjusted

**Figure 4.**
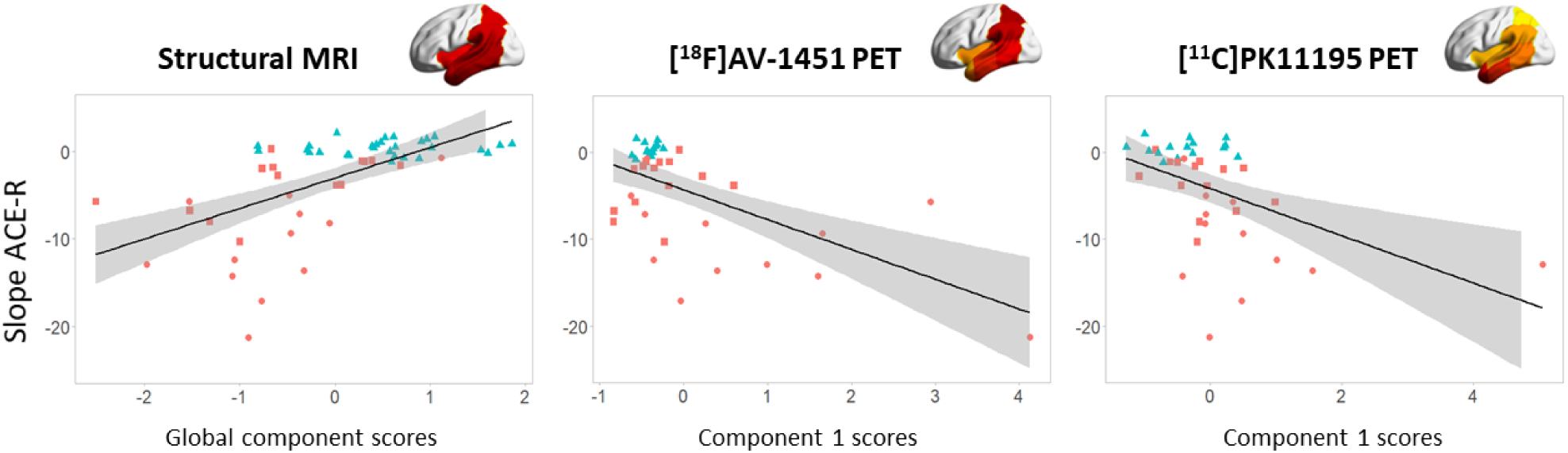
Regression analyses with annual change in scores of revised Addenbrooke’s Cognitive Examination (Slope ACE-R, y axis) and individual baseline scores for each modality-specific principal component (x axis): structural MRI (left panel), [^18^F]AV-1451 PET (middle panel), and [^11^C]PK11195 PET (right panel). Different colours represent different diagnostic groups (patients with Alzheimer’s disease = red circles, patients with amyloid-positive mild cognitive impairment = red squares, controls = blue triangles).

In patients, the final model of multiple regression on cognitive slope (adjusted R^2^ = 0.418, Std Error= 4.18; p=0.001) included both [^18^F]AV-1451 components (#1: Est=-2.57, Std Error=0.71, p=0.002; #2: Est=-1.64, Std Error=0.74, p=0.038), and the anterior [^11^C]PK11195 (#1: Est=-1.92, Std Error=0.74, p=0.017) as predictors (Fig. 5 and Table 3). Of note age, education, sex, the MRI component, and the posterior [^11^C]PK11195 component were excluded from the final model.

**Table 3.** Results of the multivariable regression models on the regression slope in patients. For both frequentist (top) and Bayesian (bottom) the estimated coefficients for variables included in the final (“best”) models are reported.

| Frequentist regression |  |  |  |  |  |  |  |  |
| --- | --- | --- | --- | --- | --- | --- | --- | --- |
| Final model<br>(Stepwise Backward selection) | Estimate | Std Error | Std Beta | t value | p | Adj R<sup>2</sup><br>(std err) | F | p |
| (Intercept) | -5.41 | 0.87 |  | -6.19 | 0.000 | 0.418<br>(4.18) | 8.05 | 0.001 |
| AV component 1 | -2.57 | 0.71 | -0.54 | -3.60 | 0.002 |  |  |  |
| AV component 2 | -1.64 | 0.74 | -0.33 | -2.21 | 0.038 |  |  |  |
| PK component 1 | -1.92 | 0.74 | -0.39 | -2.59 | 0.017 |  |  |  |
| Bayesian regression |  |  |  |  |  |  |  |  |
| Final model<br>(Bayesian Factor based selection) | Mean | Std Deviation | BF<sub>inclusion</sub> | 95% Credible interval |  | R<sup>2</sup> | BF<sub>10</sub> |  |
|  |  |  |  | Lower | Upper |  |  |  |
| (Intercept) | -6.82 | 0.82 | 1 | -8.502 | -5.129 | 0.523 | 46.56 |  |
| AV component 1 | -2.15 | 0.65 | 24.70 | -3.491 | -0.802 |  |  |  |
| AV component 2 | -1.37 | 0.68 | 4.20 | -2.774 | 0.026 |  |  |  |
| PK component 1 | -1.61 | 0.68 | 6.44 | -3.001 | -0.210 |  |  |  |

Abbreviations AV: [<sup>18</sup>F]AV-1451; PK: [<sup>11</sup>C]PK11195; BF: Bayesian factor; Std: standard; Adj: adjusted

**Figure 5.**
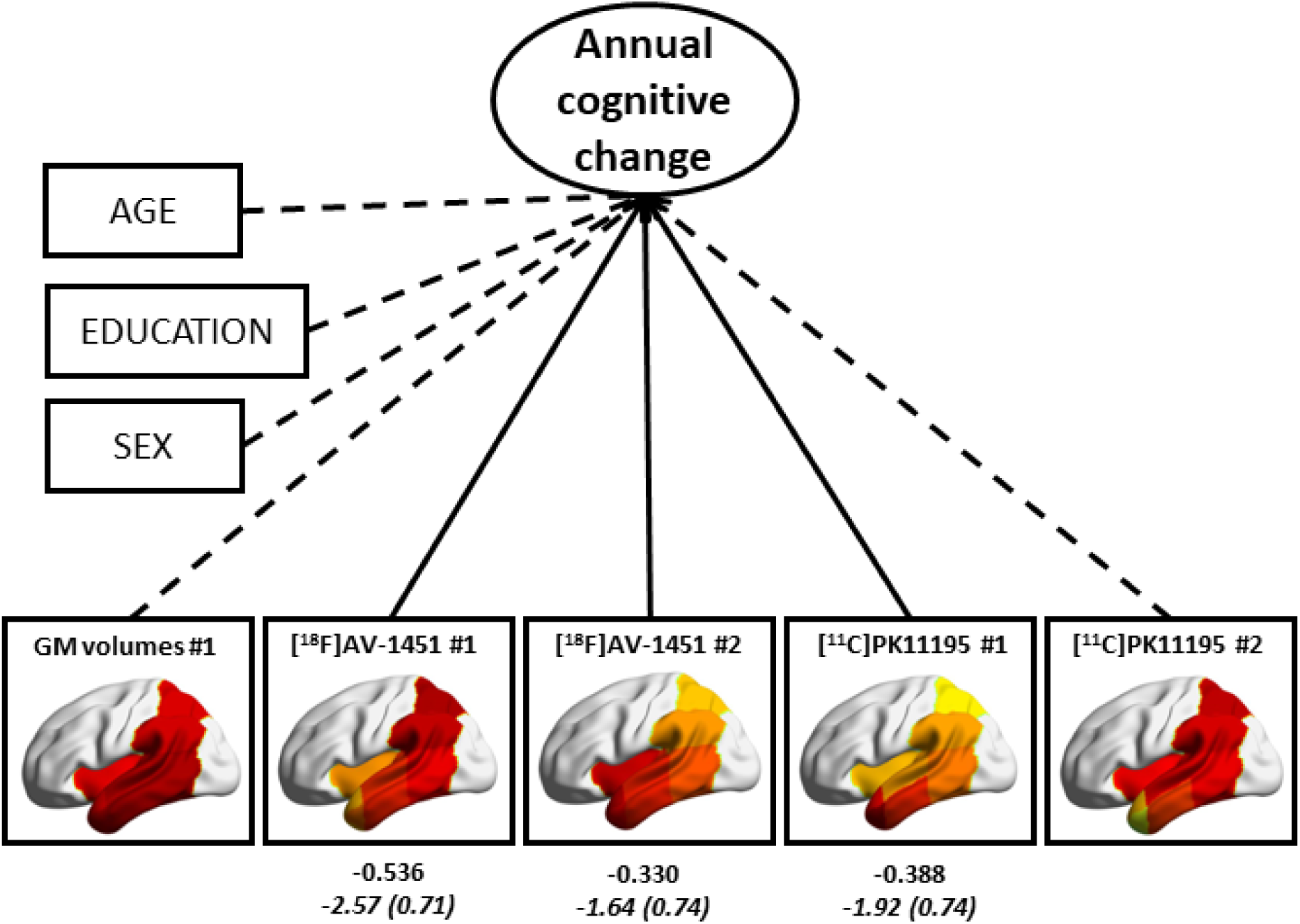
Results of the multiple linear regression in patients, with cognitive slope (annual cognitive change) extracted by the Latent Growth Curve Model as dependent variable, and brain components’ scores, age and education as independent variables. Solid arrows indicate significant coefficients of brain imaging measures indicated by the stepwise backward elimination, while dashed arrows indicate variables excluded by the final model. Values in Roman are standardized estimates, and values in italics are unstandardized beta estimates (standard errors in parentheses).

Model summary and coefficients for both the initial model (adjusted R^2^ = 0.389, Std Error= 4.43; p=0.027), the full model with only brain predictors (adjusted R^2^ = 0.474, Std Error= 4.12; p=0.002), and the final model are reported in Supplementary Table 2. Either in the initial model with covariates or in the full model with only brain measures as predictors, the posterior [^18^F]AV-1451 component and the anterior [^11^C]PK11195 component showed the highest estimated coefficients (Supplementary Table 2). In addition, the reduced multiple regression analysis, with the first component of each imaging method only, included the [^18^F]AV-1451 component (Est=-2.42, Std Error=0.77, p=0.004) and the [^11^C]PK11195 component (Est=-1.71, Std Error=0.80, p=0.042) in the final model (adjusted R^2^ = 0.366, Std Error=4.52; p=0.002), while the MRI component was discarded.

### Two-step Bayesian prediction

With all brain components and demographic variables as candidate predictors of cognitive decline, model comparison using Bayes factors indicated that the best model included both [^18^F]AV-1451 components (#1: Mean (SD) = −2.15 (0.65); BF inclusion = 24.70; #2: Mean (SD) = −1.37 (0.68); BF inclusion = 4.20), and the anterior [^11^C]PK11195 component (Mean (SD) = −1.61 (0.68); BF inclusion=6.44) as predictors (BF_10_ = 46.56; R^2^ = 0.52). Hence, the best model in this statistical framework did not contain structural MRI data. See Table 3 for details on the final model and Supplementary Table 3 for a list of models evaluated and the corresponding BF_10_. The reduced Bayesian regression analysis with only the first component of each imaging method as predictor was in accord with the frequentist approach. The best model identified with BF_10_ criteria was the one with only the posterior [^18^F]AV-1451 and the anterior [^11^C]PK11195 components only as predictors of slope (BF_10_ = 20.81; R^2^ = 0.42), not the MRI component.

## Discussion

This study demonstrates the independent and combined prognostic value of neuroimaging biomarkers for tau pathology ([^18^F]AV-1451 PET), neuroinflammation ([^11^C]PK11195 PET) and brain atrophy (structural MRI), in predicting longitudinal cognitive decline in patients with Alzheimer’s disease. Baseline markers for tau pathology, neuroinflammation and atrophy in temporo-parietal regions individually predicted cognitive decline. Faster cognitive decline was associated with higher baseline tau pathology in posterior temporo-parietal regions and increased neuroinflammation in the anterior temporal structures. The Bayesian analysis confirmed the evidence against the predictive value of MRI atrophy over and above the PET markers of tau pathology and neuroinflammation.

PCA was used to derive the most parsimonious neuroanatomical patterns of pathology that explain most of the imaging variance across the cohort. PCA indicated separate sets of regions (i.e., components) in which tau pathology and neuroinflammation clustered in anterior vs. posterior temporo-parietal regions, and this pattern is consistent with the well-established distribution of pathology in Alzheimer’s disease (Garibotto *et al*., 2017; Jagust, 2018; Whitwell, 2018). This distinction would be lost in a global average measure, while the use of separate regional values would have led to redundant multiple comparisons. In keeping with previous studies (see Chandra *et al*., 2019 and Melis *et al*., 2019 for reviews), our analyses showed that the participants’ weighting on atrophy, posterior [^18^F]AV-1451 and anterior [^11^C]PK11195 components were separately associated with more rapid cognitive decline (Fig. 4, Table 2). This result was confirmed by both the one- and two-step univariable prediction approaches. This corroborates the previously reported associations between cognitive deficits in Alzheimer’s disease and the individual effects of tau pathology, neuroinflammation, and downstream cortical atrophy (Femminella *et al*., 2016; Bejanin *et al*., 2017). Although cross-sectional imaging studies with different PET ligands have reported single associations of cognitive performance with *in vivo* tau (Brier *et al*., 2016, Cho *et al*., 2016*b*; Johnson *et al*., 2016; Ossenkoppele *et al*., 2016; Pontecorvo *et al*., 2017, see Chandra *et al*., 2019 for a review) and microglial activation (Edison *et al*., 2008; Okello *et al*., 2009, Fan *et al*., 2015*a*; Passamonti *et al*., 2018, 2019; see Chandra *et al*., 2019 for a review), less is known about their relationship to longitudinal cognitive changes. Previous PET studies in patients with Alzheimer’s dementia and MCI showed that baseline [^18^F]AV-1451 PET uptake correlates with cognitive decline over a period of six (Koychev *et al*., 2017) or 18 months (Pontecorvo *et al*., 2019). Conversely, microglial activation (as measured by [^11^C]PK11195 PET) shows progression over 14-16 months (Fan *et al*., 2015*b*, 2017), although the predictive value of baseline measures of inflammation on longitudinal cognitive decline has not been investigated yet. Other studies using [^11^C]-PBR28 to quantify neuroinflammation over a period of at least one year (median 2.7 years) in MCI and Alzheimer’s Disease reported increased microglial activation as a function of a significant worsening on the Clinical Dementia Rating scale (Kreisl *et al*., 2016). Likewise, binding of [^18^F]DPA-714, another TSPO PET ligand, is negatively associated with cognitive performance (Hamelin *et al*., 2018).

Improving our knowledge of how baseline measures of tau, neuroinflammation, and brain atrophy predict cognitive decline in Alzheimer’s disease may inform future cost-effectiveness studies in large and epidemiologically representative cohorts of patients. Although other studies have assessed the predictive value of different brain markers on longitudinal cognitive decline in Alzheimer’s disease (see Chandra *et al*., 2019 and Melis *et al*., 2019 for reviews), this study is the first one, to our knowledge, to evaluate and compare three biomarkers simultaneously (i.e., tau pathology, neuroinflammation, brain atrophy). Overall, our data are consistent with past research and highlight the added value of PET imaging over and above MRI markers. Although brain atrophy do show in isolation predictive value for cognitive decline in Alzheimer’s disease (Jack *et al*., 2015), when the models included tau burden, microglial activation and atrophy jointly, only PET was found to be predictive, but not MRI (Fig. 5, Table 3). This critical result was confirmed by both frequentist and Bayesian analyses which showed evidence against the added value of MRI data on predicting cognitive decline over and above PET assessments. This aligns with cross-sectional studies that report a stronger association of tau molecular imaging than structural MRI with cognitive performance in patients with Alzheimer’s disease (Bejanin *et al*., 2017; Mattsson *et al*., 2019). More specifically, in patients with MCI and Alzheimer’s dementia, Benjanin and colleagues reported an association between regional tau PET binding and cognitive impairment, which was partly mediated by grey matter volumes (Bejanin *et al*., 2017). Cognition was equally explained by brain atrophy and tau pathology, but after accounting for grey-matter values, *in vivo* tau pathology remained correlated with cognitive performance (Bejanin *et al*., 2017). Likewise, Mattson et al. (2019) found that both [^18^F]AV-1451 PET and structural brain MRI are associated with cognition in Alzheimer’s disease (spanning preclinical, prodromal, and dementia stages), although associations of tau PET indices were stronger than those for MRI markers (Mattsson *et al*., 2019).

Our data suggest that posterior temporo-parietal [^18^F]AV-1451 binding and anterior temporal [^11^C]PK11195 binding were associated with cognitive decline. In patients with Alzheimer’s disease, temporo-parietal cortical tau PET signal is consistent with Braak stage III and above, while in cognitively healthy older people, the signal is localised to entorhinal cortex and inferior temporal cortex (Cho *et al*., 2016*a*; Johnson *et al*., 2016; see Jagust, 2018 for a review). *Post mortem* studies have likewise reported tau deposition in the medial temporal cortex in healthy elderly people and Alzheimer’s dementia (Jagust, 2018). Tau burden in the entorhinal, limbic, and temporal neocortex relates to cortical atrophy in patients with MCI and Alzheimer’s disease, although not in cognitively normal controls (Timmers *et al*., 2019). These findings suggest that tauopathy in the medial part of the temporal lobe may be an age-related norm, rather than indicative of Alzheimer’s disease cognitive decline (Femminella *et al*., 2018). For this reason, tau PET binding here may be a weaker predictor for cognitive decline than tau in the posterior temporo-parietal regions. The co-occurrence with amyloid-β and neuroinflammation may induce the tau spreading from the medial temporal lobe to other cortical regions, which may be associated with downstream neurodegenerative processes and cognitive decline (Mhatre *et al*., 2015; Jagust, 2018; Perea *et al*., 2018). This suggests a driving role of neuroinflammation in tau spread and neurodegeneration in Alzheimer’s disease (Yoshiyama *et al*., 2007; Asai *et al*., 2015; Maphis *et al*., 2015), in which activated microglia facilitate tau spread (Maphis et al., 2015; Perea et al. 2018).

There are limitations to our study. [^11^C]PK11195 PET is not only tracer for microglial activation. Several second-generation PET radioligands for TSPO have been developed since [^11^C]PK11195 (e.g., [^11^C]PBR28 and [^18^F]DPA-714), and used in human studies (Vivash and OBrien, 2016). They are characterised by higher signal-noise ratio and lower lipophilicity than [^11^C]PK11195. However, they require genetic analysis to assess a single-nucleotide polymorphism (rs6971), which influences their binding affinity and causes heterogeneity in PET data (Dupont *et al*., 2017). In contrast, [^11^C]PK11195 is not significantly affected by this polymorphism and hence can be used across the whole patient population, making it our preferred method to study microglia activation. Second, the cross-sectional nature of our imaging assessment did not enable mediation analysis of the causality between tau pathology, microglial activation progression and their effect on cognitive decline. However, we speculate that both processes are directly involved in mediating the rate of cognitive deterioration in Alzheimer’s disease, carrying some unique information across the disease spectrum. Third, the modest sample size of our cohort limited the applicability of the one-step prediction procedure with multiple predictors, which may lead to a more precise prediction than the two-step procedures. However, both frequentist and Bayesian multivariable approaches give similar results, aligning with those obtained by the one-step prediction. The convergence between all statistical models (i.e., LGCM with predictors, linear regression and Bayesian model) mitigates against sample-dependant biases on the estimation of the most parsimonious model.

We conclude that the PET markers of regional pathological processes are stronger predictors than atrophy as measured by MRI. The models were convergent in identifying tau burden in posterior cortical regions and neuroinflammation in the anterior temporal lobe as key imaging predictors of cognitive decline in the clinical spectrum of Alzheimer’s disease. In contrast, atrophy measures predicted cognitive decline only if considered individually but not over and above the effects of tau burden and inflammation when we consider all processes together. Our findings support the use of PET imaging of tau pathology and microglial activation for prognostication and patients’ stratification in clinical trials.

## Acknowledgments

We thank our participant volunteers for their participation in this study, and the radiographers/technologists at the Wolfson Brain Imaging Centre and Addenbrooke’s PET/CT Unit, and the research nurses of the Cambridge Centre for Frontotemporal Dementia and Related Disorders for their invaluable support in data acquisition. We thank the East Anglia Dementias and Neurodegenerative Diseases Research Network (DeNDRoN) for help with subject recruitment, and Drs Istvan Boros, Joong-Hyun Chun, and other WBIC RPU staff for the manufacture of the radioligands. We thank Avid (Lilly) for supplying the precursor for the production of [^18^F]AV-1451 used in this study.

